# On-device edge-learning for cardiac abnormality detection using a bio-inspired and spiking shallow network

**DOI:** 10.1101/2023.12.15.23299994

**Authors:** Zhaojing Huang, Wing Hang Leung, Leping Yu, Luis Fernando Herbozo Contreras, Ziyao Zhang, Nhan Duy Truong, Armin Nikpour, Omid Kavehei

**Affiliations:** School of Biomedical Engineering at The University of Sydney, Australia; Central Clinical School of the University of Sydney, Australia

**Keywords:** Spiking neural networks, Electrocardiogram analysis, Energy efficiency, On-device fine-tuning, Generalization, Robustness

## Abstract

This work presents an on-device edge-learning for cardiac abnormality detection by developing a hybrid and spiking form of 2-Dimensional (time-frequency) Convolutional Long-Short-Term Memory (ConvLSTM2D) with Closed-form Continuous-time (CfC) neural network (sCCfC), which is a bio-inspired shallow network. The model achieves an F1 score and AUROC of 0.82 and 0.91 in cardiac abnormalities detection. These results are comparable to the non-spiking ConvLSTM2D-CfC (ConvCfC) model^1^. Notably, the sCCfC model demonstrates a significantly higher energy efficiency with an estimated power consumption of 4.68 *µ*J/Inf (per inference) on an emulated Loihi’s neuromorphic chip architecture, in contrast to ConvCfC model’s consumption of 450 *µ*J/Inf on a conventional processor. Additionally, as a proof-of-concept, we deployed the sCCfC model on the conventional and relatively resource-constrained Radxa Zero, which is equipped with Amlogic S905Y2 processor for *on-device training*, which resulted in performance improvements. After initial training of 2 epochs on a conventional GPU, the F1 score and AUROC improved from 0.46 and 0.65 to 0.56 and 0.73 respectively with 5 additional epochs of on-device training. Furthermore, when presented with a new dataset, the sCCfC model showcases strong out-of-sample generalization capabilities that can constitute a pseudo-perspective test, achieving an F1 score and AUROC of 0.71 and 0.86. The spiking sCCfC also outperforms the non-spiking ConvCfC model in robustness regarding effectively handling missing ECG channels during inference. The model’s efficacy extends to single-lead electrocardiogram (ECG) analysis, demonstrating reasonable accuracy in this context, while the focus of our work has been on the computational and memory complexities of the model.

## 1. INTRODUCTION

Many technologies have been created to track heart activity, and because it is non-invasive and reasonably priced, the Electrocardiogram (ECG) has become a popular option. The 12-lead ECG is considered the gold standard for assessing cardiac electrical activity in clinical settings^2^. This research introduces a Spiking Neural Network (SNN) that incorporates a bio-inspired Neural Circuit Policy (NCP)^3^ based model developed on 2-Dimensional (time-frequency)^4^ Convolutional Long-Short-Term Memory (ConvLSTM2D)^5^. This model is explicitly designed for abnormality detections where large models cannot be deployed and achieves an average F1 score of 0.82. In contrast to conventional artificial neural networks, SNNs exhibit significantly lower energy consumption while maintaining a reasonable performance, primarily due to their ability to generate sparse output spike trains when the integrated input surpasses a predetermined threshold^6^. The aim of developing an SNN architecture in this work was to demonstrate a feasibility study for a significant reduction in power consumption of the ECG abnormality detection system, opening new avenues for edge-AI and near-sensory computing in medical devices. Therefore, the mere choice of a 12-lead ECG is for that purpose only, and this method can be expanded to other domains, including single-lead ECGs and implantable cardiac monitoring devices (ICMs).

Considering the promise of SNNs in enabling highly power-efficient smart systems, this research is influenced by the progress made in bio-inspired Neural Circuit Policy (NCP) models, which are designed to aid healthcare professionals in detecting cardiac abnormalities^1^. Unlike traditional Recurrent Neural Networks (RNNs) such as Long Short-Term Memory (LSTM), these NCP structured models effectively address the complexities inherent in learning long-term dependencies within specialized tasks, as expounded upon in the investigation conducted by Lechner *et al*. ^3^. Furthermore, the research by Lechner *et al*. ^3^ substantiates the computational superiorty of the NCP model when compared to contemporary deep learning models.

Unlike traditional deep neural network models that heavily depend on clean input data, the NCP model displays enhanced robustness when faced with transient disruptions commonly encountered in real-world scenarios^3^. Moreover, the NCP model’s simplified and sparse network structure facilitates easier interpretation^3^. Furthermore, its minimal memory requirements render it well-suited for deployment on various hardware platforms^1^. An important feature of the NCP model is its capacity to perform effectively with a relatively modest number of neurons. The present study’s SNN model achieved an F1 accuracy score of 0.82. The envisioned application of this research, as depicted in Figure 1, holds the potential for expansion onto hardware, thus enabling seamless integration into wearable devices.

**FIG. 1.**
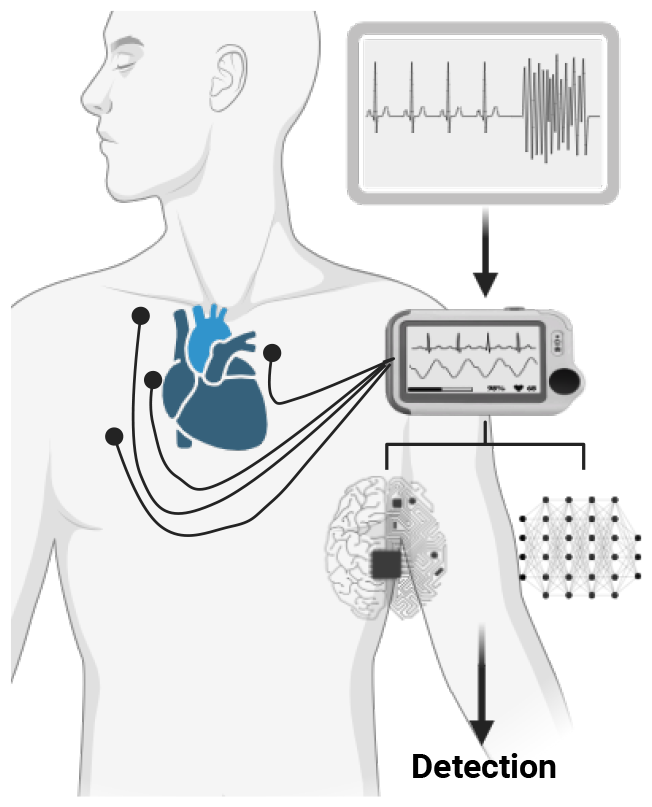
The proposed model and use for wearable devices. On-device and real-time detection and identification of abnormalities, harnessing the potential of powerful neuromorphic chips and spiking neural networks.

In the current study, on-chip learning was also conducted, demonstrating that the current model can undergo fine-tuning directly on the chip. This allows the model to be customized based on a patient’s specific data, enhancing its ability to generalize across different patients. The on-chip tuning improves the F1 score and AUROC from 0.31 and 0.63 to 0.45 and 0.72 when training with 640 samples is performed.

### A. Background

Medical diagnostics has witnessed substantial advancements in abnormality detection owing to the introduction of state-of-the-art models. For instance, Petmezas *et al*. ^7^ intrduced a Hybrid CNN-LSTM Network tailored to classify specific heartbeat types using ECG data, delivering exceptional sensitivity (97.87%) and specificity (99.29%). On another front, Chen *et al*. ^8^ engineered a model comprising five CNN blocks, a bidirectional RNN layer, an attention layer, and a dense layer, culminating in an impressive F1 score of 0.84 for detecting various ECG abnormalities. Huang *et al*. ^9^ took a rapid approach, introducing a compression residual convolutional neural network-based model designed for ECG classification, which achieved an average accuracy of 98.79%. Furthermore, Huang *et al*. ^10^ unveiled a shallow S4D model, which demonstrated remarkable robustness with an F1 score of 0.81 and a remarkable capability to manage situations involving incomplete data inputs.

Shifting our focus to innovations in the NCP model domain, Mathias Lechner’s NCP model, initially devised for car driving applications employing camera input, stands out for its unique emphasis on the road’s horizon, a departure from the conventional CNN models that typically prioritize roadside features. The NCP model’s dedicated attention to the road’s horizon equips it to capture and learn global driving features adeptly. This prowess is further evidenced by the remarkable variance explained by the first principal component (PC1), reaching an impressive 92%^3^. Such proficiency in comprehending global driving features holds significant promise for enhancing decision-making in car driving applications. Additionally, the field has seen advancements from Ramin Hasani, who has pushed the boundaries of Liquid Time-constant Networks by developing closed-form continuous-time neural networks, ushering in improved computational speed and efficiency^11^. Recently, Huang *et al*. ^1^ has developed an NCP-based model for cardiac abnormality detection.

Like bio-inspired NCP models, spiking neural networks have found their application in cardiology. This utilization of SNNs showcases their potential to reduce power consumption and underscores their applicability in addressing realworld challenges such as ECG classification. Yan, Zhou, and Wong ^12^ developed an SNN for a portable device to classify ECG beats and achieve better energy efficiency and enables a daily monitoring and classifying of ECG. Rana and Kim ^13^ developed spike-timing-dependent plasticity (STDP) for ECG classification. The model weights are trained according to spike signal timing and reward or punishment signals to save power on a portable device. Feng *et al*. ^14^ developed an artificial neural network for ECG classification and transferred into spiking.

## II. PREREQUISITE

### A. Spiking Neural Network (SNN)

Spiking Neural Networks (SNNs) are similar to Artificial Neural Networks (ANNs) in that they utilize computing units with continuous activation values and a set of weighted inputs, as described by Tavanaei *et al*. ^6^. In this SNN architecture, one finds interconnected spiking neurons connected by synapses characterized by their adjustable scalar weights. It’s important to note that SNNs encompass two distinct types of synapses: excitatory synapses, which lead to an increase in the membrane potential upon receiving input, and inhibitory synapses, which induce a decrease in the membrane potential upon stimulation^6^. The generation of a spike in an SNN depends on the cumulative effect of stimulus changes surpassing a certain threshold. This feature contributes to the network’s energy-efficient properties, as explained in the same source^6^. The implementation of the SNN in this research was carried out utilizing the “snnTorch” package^15^.

### B. Neural circuit policies (NCP)

The NCP model, as introduced by Lechner and colleagues in their research^3^, constitutes an end-to-end learning system characterized by convolutional layers. This model draws its foundational inspiration from the intricate neural wiring diagram of the C. elegans nematode, as extensively detailed in Lechner *et al*. ^3^. Within the biologically inspired framework of the NCP model, four distinct neural layers are at play: sensory neurons (*N*_*s*_), interneurons (*N*_*i*_), command neurons (*N*_*c*_), and motor neurons (*N*_*m*_). A specific number of synapses are strategically introduced to facilitate the seamless flow of information between each consecutive layer, thereby governing the synaptic connectivity.

A comprehensive exposition of the fundamental aspects of the NCP network and its specific details can be found in the supplementary information accompanying this manuscript.

### C. Closed-form Continuous-time (CfC) Neural Networks

Closed-form Continuous-time (CfC) Neural Networks, as innovatively presented by Hasani and collaborators^16^, distinctly depart from traditional practices by obviating the need for numerical solvers when generating temporal rollouts within the realm of Ordinary Differential Equations (ODEs). These networks seamlessly integrate the strengths of ODE-based counterparts, encompassing flexibility, causality, and continuous-time attributes, all while delivering a marked enhancement in computational efficiency^16^. Equation 1 vividly encapsulates the essence of the CfC model, where the continuous-time gating mechanisms, denoted by *σ* (*−f* (*x, I*; *θ*_*f*_)*t*) and [1*−σ* (*−*[*f* (*x, I*; *θ*_*f*_)]*t*)], underpin the model’s innovative approach^16^.

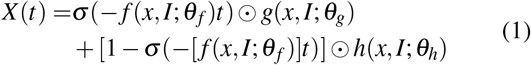

### D) ConvLSTM2D CfC (ConvCfC)

The ConvLSTM2D CfC (ConvCfC) model is structured with a single ConvLSTM2D layer responsible for feature extraction. This layer is connected to 75 neurons, which act as input neurons for the NCP (Neural Control Policy) network within the CfC arrangement^1^. The ConvCfC model, known for its compact and efficient design, demonstrated successful deployment on the STM32F746G Discovery board, an edge device characterized by resource constraints. It efficiently conducted inferences on ECG data during the deployment.^1^

## III. DATASET

In this research, the same datasets were employed for training and evaluation as the ConvCfC proposed by Huang *et al*. ^1^. The training dataset originated from the Telehealth Network of Minas Gerais (TNMG) dataset. A balanced subset of the TNMG dataset encompassing six distinct cardiac abnormalities: Atrial Fibrillation (AF), Left Bundle Branch Block (LBBB), First Degree Atrioventricular Block (1dAVb), Right Bundle Branch Block (RBBB), Sinus Tachycardia (ST), and Sinus Bradycardia (SB) was deployed in training the sCCfC. These ECG recordings are uniformly sampled at a rate of 400 Hz.

For assessing model generalization, we employed the China Physiological Signal Challenge 2018 (CPSC) dataset, which includes eight different abnormalities, with four of them overlapping with the TNMG dataset: Atrial Fibrillation (AF), Right Bundle Branch Block (RBBB), First Degree Atrioventricular Block (1dAVb), and Left Bundle Branch Block (LBBB). It’s important to note that the CPSC dataset comprises 12-lead electrocardiogram (ECG) recordings sampled at a uniform rate of 500 Hz. To ensure compatibility with the TNMG dataset used for model training, we performed a down-sampling process to harmonize the sampling rate of the CPSC dataset with that of the TNMG dataset.

For more detailed information about the dataset, please refer to the supplementary information provided at the end of this study, which offers a comprehensive overview of the composition and characteristics of both the TNMG subset and CPSC datasets used in our research.

## IV. METHODS

Our primary aim is to develop a compact and efficient model optimized for processing electrocardiogram (ECG) data, strongly emphasising energy efficiency. To achieve this objective, we have adopted the ConvCfC model, as outlined in^1^, as the foundational framework for our work. Incorporating spiking neural networks, well-known for their energy-efficient properties, is a central focus of our research. Therefore, our primary efforts are to seamlessly integrate spiking components into the ConvCfC model to attain the desired energy-efficient results.

Furthermore, we comprehensively assess the recently integrated model’s performance. This evaluation considers its capacity for generalization and its resilience in dealing with incomplete data, comparing it to the ConvCfC model as a reference point.

With the ConvCfC model, we undertake crucial preprocessing steps on the ECG data, involving filtering and Short-Time Fourier Transform (STFT) transformation. These procedures are vital for improving data quality and facilitating the extraction of relevant features.

Following preprocessing, the refined data is input into the proposed models for both training and validation. We employ selected metrics to evaluate the model’s efficacy, providing valuable insights into its performance and capabilities.

### A. Preparatory Steps for ECG Data

A Butterworth band-pass filter is strategically employed in the signal processing pipeline to attenuate the interference caused by extraneous noise within the electrocardiogram (ECG) signal. The choice of the Butterworth filter is predicated on its ability to maintain a consistent response across the entire spectrum of desired frequencies, as supported by reputable sources^17^. This band-pass filter’s passband is thoughtfully set to span from 0.5 Hz to 40 Hz. This particular frequency range is deliberately selected to retain and safeguard critical components within the ECG signal, including but not limited to the T wave, P wave, and QRS complex. Concurrently, this setting effectively obviates the presence of powerline noise at the 50 Hz frequency^18^.

After the application of the aforementioned filtration procedure, STFT is enlisted as the analytical tool of choice. Diverging from the Fast Fourier Transform (FFT), which conducts a holistic spectral analysis of the entire signal, the STFT adopts a segmented approach. It subdivides the signal into smaller, overlapping time windows, conducting FFT analysis independently on each of these temporal segments. This methodological divergence facilitates the extraction of both frequency and time-related attributes from the signal’s composition. Under this temporal segmentation, the STFT captures nuanced variations in the signal’s characteristics over time, engendering a more intricate and precise representation of the signal’s time-dependent frequency components. Consequently, this analytical approach affords comprehensive insights on both frequency-based and temporal aspects of the signal’s characteristics.

In addition to the preprocessing steps detailed in ConvCfC^1^, this study incorporates two supplementary procedures. Firstly, the preprocessed data undergoes a scaling process involving the application of the max-min scaling technique to each data point. This particular step is informed by prior research^19^ and enhances the overall data quality. Furthermore, the processed electrocardiogram (ECG) data is subjected to a spike generation process. This operation renders the data compatible with input requirements for a spiking neural network.

### B. Model ConvCfC and NCP

Taking inspiration from the Caenorhabditis elegans nematode, neural circuit policies (NCPs) have emerged as intelligent agents influenced by brain-inspired principles. These NCPs incorporate neurons with enhanced computational capabilities, resulting in the creation of sparse networks, as detailed in^3^. These networks include Liquid Time Constant (LTC) neurons, depicted as leaky integrators, which accumulate and release charge overtime to process temporal information^3^.

Furthermore, this study introduces Closed-form Continuous-time Neural Networks (CfC) for time-series modeling. Derived from liquid networks, CfC models employ closed-form ordinary differential equations (ODEs) to approximate solutions for previously unsolved integrals, significantly enhancing their performance compared to advanced, recurrent neural networks, as elaborated in^16^. The ConvCfC model, introduced in the reference^1^, is a straightforward model architecture that leverages the principles of NCP. This model is designed with simplicity and effectiveness and aims to provide an efficient approach to solving specific tasks or problems.

### C) Simple Spiking Model Design

The proposed model architecture, as illustrated in Figure 2, is built upon the foundation of the ConvCfC model introduced in the reference^1^. This model implements a streamlined architecture, featuring a singular SConv2dLSTM layer responsible for spiking feature extraction. This layer seamlessly connects to a dense layer composed of 75 LIF (Leaky Integrate-and-Fire) neurons, which function as the input neurons for the NCP network. By adopting the ConvCfC framework, the resulting model falls under the category of sCCfC models, which include 14 interneurons and command neurons, along with 6 LIF output neurons.

**FIG. 2.**
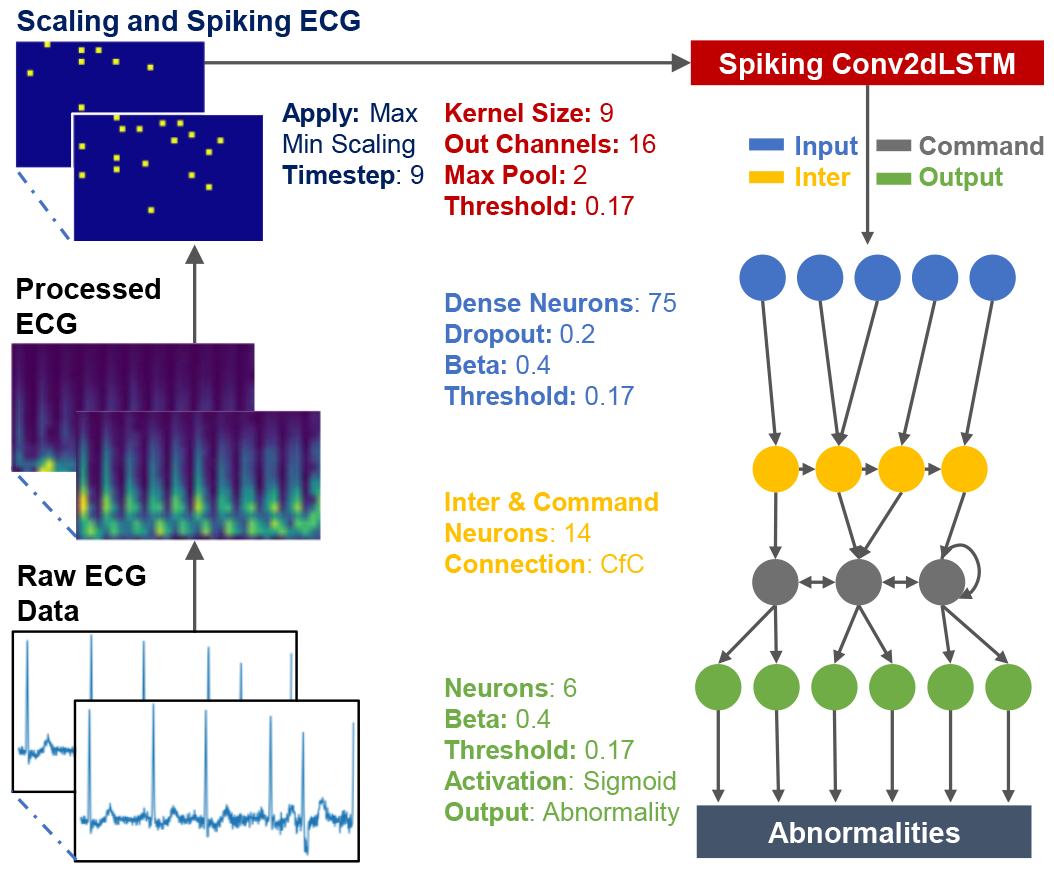
The model architecture includes a specialized Spiking SConv2dLSTM layer for processing spiking STFT-transformed ECG data. This layer extracts key features, which are then fed into 75 Leaky Integrate-and-Fire (LIF) neurons, forming the input layer for the Neuronal Coherence Propagation (NCP) network. The network’s connections are facilitated through CfC network, with a sigmoid activation function applied to the LIF neurons in the final stage.

### D. Performance Metrics

Recall and precision are pivotal metrics for appraising model performance. Precision scrutinizes the precision of positive predictions, whereas recall quantifies the proportion of actual positives correctly identified. The F1-score, a harmonious measure, amalgamates precision and recall, furnishing valuable insights into the equilibrium between the two. In this study, a threshold of 0.5 is employed for performance metrics.

The Area Under the Receiver Operating Characteristic curve (AUROC) scrutinizes a model’s capacity to discriminate between negative and positive cases across diverse threshold values. In binary classification tasks, AUROC stands as a valuable metric of assessment.

These metrics, encompassing the F1-score, precision, recall, and AUROC, are critical in thoroughly evaluating machine learning model performance, particularly in endeavors such as abnormality detection.

## V. EXPERIMENTS

### A. Train

In the experimental phase, we employed the preprocessed TNMG subset dataset as the foundation for training and validation procedures concerning the sCCfC model. To address the inherent issue of class imbalance within multi-labeled datasets, we introduced the utilization of class weights. Specifically, a weight factor of 6 was incorporated into the construction of the loss function, to afford greater consideration to positive-labeled data points. This approach aimed to rectify the imbalance between positive and negative labels in the training dataset.

The model training process was executed on a Tesla V100S-PCIE-32GB, utilizing a batch size of 32 and spanning 100 training epochs. A learning rate of 0.0003 was adopted, and optimization was performed using the AdamW optimizer in conjunction with the binary cross-entropy loss function.

To assess the models’ generalization capabilities, we turned to the preprocessed CPSC dataset. Furthermore, we evaluated the models’ robustness by subjecting them to corrupted data inputs, thereby scrutinizing their performance under challenging conditions.

### B. Model Training and Validation using In-sample Data

To assess the model’s performance, we set aside a dedicated validation subset comprising 20% of the TNMG dataset. Importantly, these validation data points were excluded from the model’s training process, exclusively for performance evaluation. This separation between training and validation was rigorously upheld for a consistent and unbiased assessment of the ConvCfC model. This meticulous validation ensures that our evaluation remains aligned with the model’s training process, enabling a robust comparison of its performance.

### C. Energy Usage of the Model

The model’s energy consumption was assessed using the *KerasSpiking* library, which offers a tool to estimate the model’s energy consumption under different deployment scenarios, such as on a neuromorphic chip or for inference on a CPU. This tool served as a means to compare the differences in energy consumption between the sCCfC and ConvCfC models.

### D. Radxa Zero Edge AI Deployment

To demonstrate the feasibility of our concept and as an alternative to employing a neuromorphic chip, we implemented the sCCfC model on a conventional computing but relatively resource-constraint system, Radxa Zero, which is a single-board computer, featuring an Amlogic 905Y2 processor with 64-bit ARM architecture and up to 4GB of 32-bit memory. This could appropriately emulate what is possible if a digital silicon chip with custom computing architecture is designed.

The experimental model refinement process was conducted on the Radxa Zero. Initially, a model pre-trained for two epochs on the TNMG dataset using a GPU was chosen as the foundational model for fine-tuning. Adjustments were made to accommodate the limited computational resources available on the board, including reducing the batch size from 32 to 8 during model training on the GPU. A dataset comprising 640 data points was employed for fine-tuning throughout five epochs, with subsequent validation conducted on a subset of 320 data points. Following the fine-tuning process, the model was inferred on a data set containing 128 data points, and the results were compared to those obtained from the base model originally trained on the GPU for two epochs.

### E. Assessment of Data on Unfamiliar Samples

The performance assessment of the models will be conducted meticulously using the CPSC dataset, thoughtfully curated to mirror real-world scenarios. Through a rigorous comparative analysis of the models’ predictions vis-à-vis established ground truth values, we intend to discern their respective merits and demerits. This empirical evaluation will provide valuable insights into the efficacy of the models and may offer suggestions for potential enhancements in the realm of ECG analysis. A critical comparative analysis of the performance of sCCfC and ConvCfC models will be conducted.

### F. Model Robustness

In this section, we will subject the model to a rigorous battery of robustness tests. These tests involve systematically removing random channels from the 12-lead ECG data. The primary objective is to assess how well the model performs when confronted with missing input information. We will progressively remove varying numbers of channels, ranging from 1 to 6 leads, in order to evaluate the model’s ability to maintain accuracy across different degrees of data incompleteness.

To quantify and compare the model’s performance in these diverse scenarios, we will employ a set of performance metrics. These metrics will provide valuable insights into potential areas for model improvement and offer a deeper understanding of possible refinements. Furthermore, we will compare the performance of the sCCfC model with its non-spiking counterpart, the ConvCfC model^1^, to shed light on how spiking dynamics impact the model’s robustness.

### G. Model Evaluation: Single Lead ECG Processing

In our extensive study, we conducted a thorough analysis to assess the model’s proficiency in handling Single Lead ECG, specifically emphasizing Lead II. The model underwent specialized training, utilizing only Lead II data from the dataset.

We then exclusively evaluated its performance with Single Lead ECG, comparing it to the model trained on the complete 12-lead ECG dataset. It’s noteworthy that the training process and model architecture remained consistent with the 12-lead examination. The primary distinction lies in integrating Lead II ECG data for both training and testing, providing precise insights into the model’s capabilities within this specific context.

## VI. RESULTS

In this section, we will perform an exhaustive analysis of the model’s performance, delving deeply into their capabilities and resilience.

### A. Model Performance

This study primarily concentrates on the training phase of the proposed models, as elaborated upon in preceding sections of this paper. Specifically, we employ the TNMG subset dataset, as previously detailed, as the primary dataset for training and evaluating the models under investigation. The training procedure encompasses 100 epochs, during which we meticulously track and document key metrics such as accuracy and loss for both the validation and training datasets at each epoch. Visual representations in this scholarly work provide a clear graphical overview of these recorded metrics, facilitating a comprehensive understanding of the entire training process. Additionally, we conduct a thorough assessment of the sCCfC model’s performance, and the results of this evaluation are visually presented in Figure 3 for ease of reference.

**FIG. 3.**
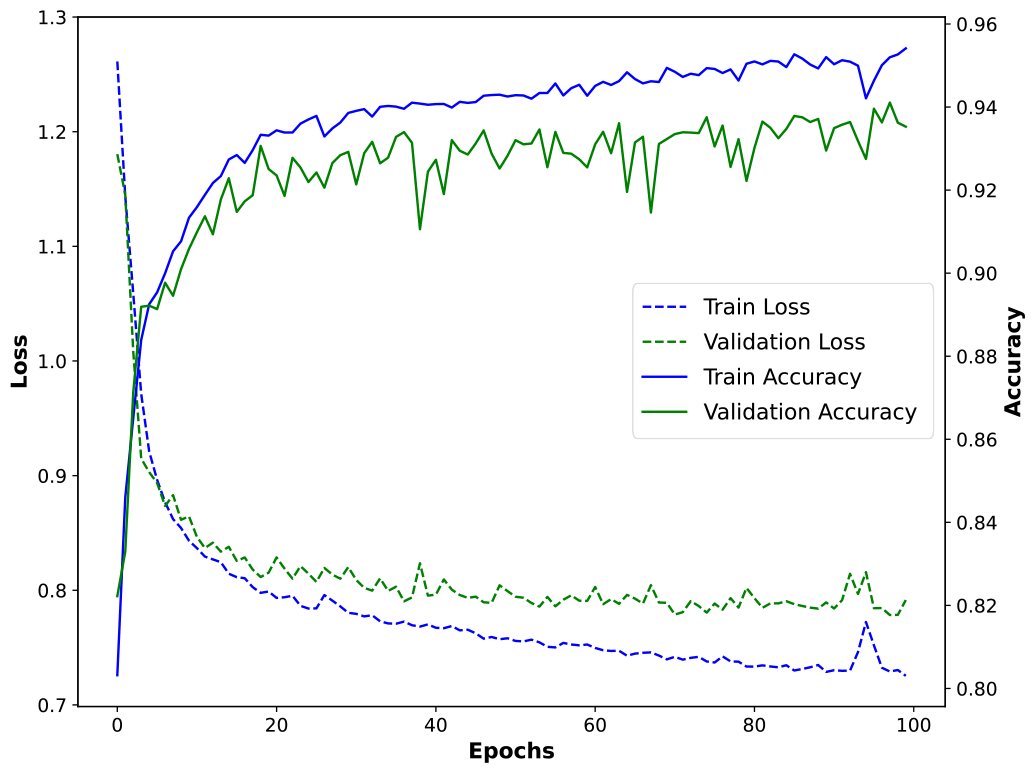
The figure provides a visual representation of the model’s performance throughout the training process, displaying both training and validation accuracy and losses.

Figure 3 reveals a prominent pattern as the training process unfolds. Notably, there is a noticeable decrease in training loss, indicating continuous improvement in the model’s performance. Simultaneously, training accuracy steadily increases and eventually stabilizes, reflecting the model’s effective learning from the training data.

Throughout the training duration, a consistent trend emerges in the validation accuracy, with a steady rise indicating enhanced model performance on unseen data. Furthermore, validation loss consistently decreases over time, suggesting that the model’s predictions increasingly align with the actual labels during the validation phase.

The evaluation of the sCCfC models’ performance is succinctly summarized in Table I, with ConvCfC’s performance serving as a baseline for comparison in Figure 4. Comparing the two models, the sCCfC model has achieved slightly lower average F1 score and AUROC values in in-sample validation, specifically recording 0.82 and 0.91 compared to ConvCfC’s 0.83 and 0.96, respectively. However, it’s crucial to emphasize that the decrease in performance is minimal, especially when considering the F1 score.

**FIG. 4.**
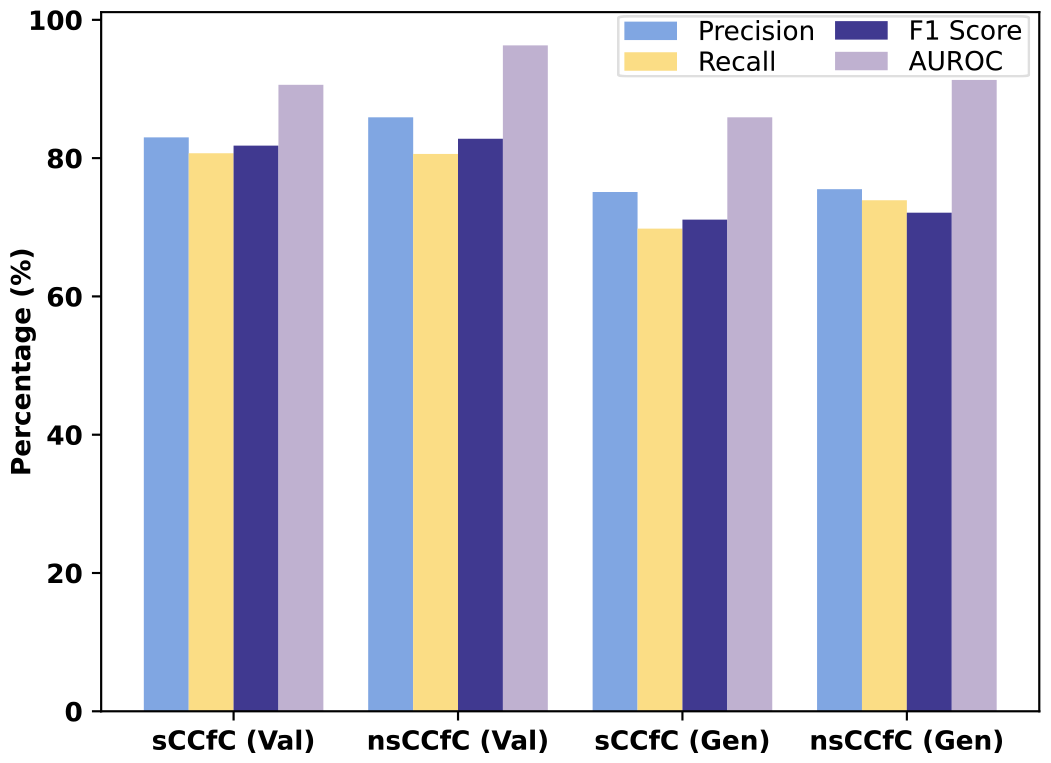
The sCCfC model’s comprehensive performance, both insample and during generalization, is benchmarked against the ConvCfC model^1^, serving as a foundational reference.

**TABLE I.**
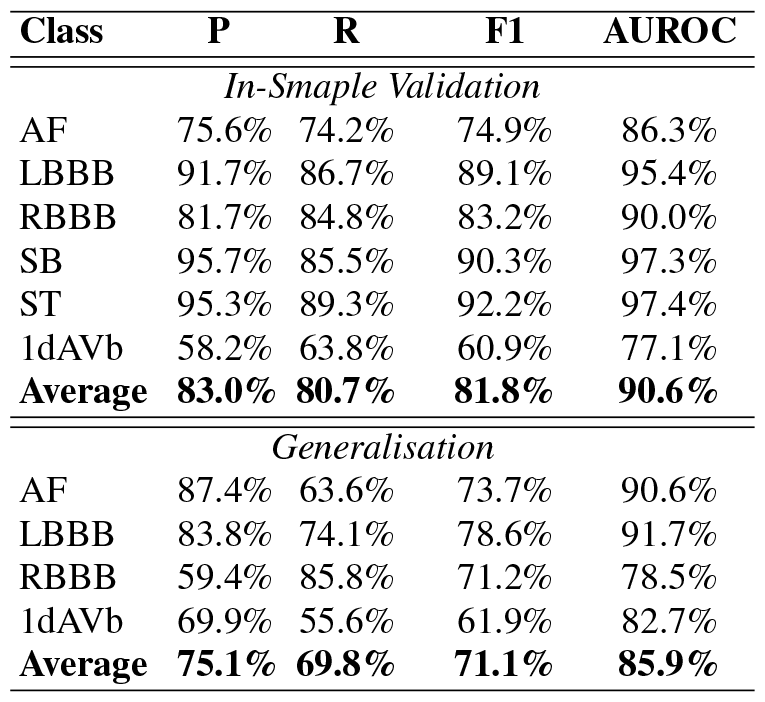
In-sample Validation and Generalisation Results for the sCCfC Model (P: Precision, R: Recall)

It becomes apparent that the sCCfC model’s performance closely mirrors that of ConvCfC. This observation underscores the robust performance of sCCfC, even when operating in spiking settings.

While accuracy is a common metric for evaluating model performance, it may not always provide a comprehensive assessment. Nevertheless, it’s worth noting that during the training process, we recorded validation accuracy metrics. Interestingly, in the validation phase, the sCCfC model achieved an accuracy of 94.1%, demonstrating its strong performance. However, it’s noteworthy that this accuracy was only marginally lower than the 94.8% achieved by the ConvCfC model. This minor difference suggests that while the sCCfC model was expected to perform slightly worse than the ConvCfC model, the actual gap in performance turned out to be quite small.

### B. Energy Usage of the Model

Energy consumption assessments for both the sCCfC and ConvCfC models were conducted utilizing the KerasSpiking framework. Specifically, the ConvCfC model demonstrated an estimated energy consumption of approximately 450 *µ*J/Inf (CPU), whereas the sCCfC model displayed notably lower energy consumption, estimated at approximately 4.68 *µ*J/Inf (Loihi). A comprehensive breakdown of these energy consumption estimates can be found in Table II.

**TABLE II.** Energy Consumption per Inference for Each Layer.

| Layer | CPU ($\mu\text{J}$) | Loihi ($\mu\text{J}$) |
| --- | --- | --- |
| ConvLSTM2D | 110 | 4.6 |
| Dense | 340 | 0.055 |
| CfC | 0.1 | 0.0044 |
| Sigmoid | 0.052 | 0.0044 |

### C. On-Device Fine-Tuning

This study implemented the sCCfC model on the Radxa Zero with an Amlogic 905Y2 processor and 4GB memory as fully digital and conventional computing on a small board alternative to a neuromorphic chip. Model refinement on the Radxa Zero involved using a pre-trained model, originally trained for two epochs on a large dataset (TNMG), via a GPU. Adjustments were made, such as reducing the batch size to 8, during fine-tuning with 72% memory utilization. The fine-tuning process included training on a dataset of 640 data points over five epochs and validation on a subset of 320 data points. The performance of the fine-tuned model was evaluated by comparing its results to those of the GPU-trained base model.

Throughout the training process, the model exhibited notable enhancements in performance metrics. Specifically, in the 320-sample validation dataset, the average F1 score increased from 0.46 to 0.56, while the AUROC improved from 0.65 to 0.73. Additionally, when testing the fine-tuned model on a larger test set of 1280 samples, the average F1 score and AUROC showed substantial improvements, rising from 0.31 and 0.63 in the pre-trained model to 0.45 and 0.72.

This observation underscores the effectiveness of on-device fine-tuning in enhancing model performance. By conducting the fine-tuning process directly on the hardware, in this case, the Radxa Zero, we can clearly see tangible evidence of its positive impact. On-device fine-tuning is a testament to the adaptability and resource efficiency of the sCCfC model. It signifies that the model is capable of initial training and further refinement within the constrained computational environment of the Radxa Zero.

### D. Model Generalization

Our evaluation involved applying predictive modeling to the CPSC dataset using models that had previously undergone training on the TNMG subset, as elaborated in the data section. The primary objective of this assessment was to gauge the models’ capacity for generalization, especially when confronted with new and unfamiliar data from the CPSC dataset, which significantly differs from their original training data.

It’s crucial to emphasize that the CPSC dataset comprises eight distinct types of abnormalities, with only four overlapping those in the TNMG dataset. Evaluating the model’s performance on this subset provided valuable insights into its ability to transfer acquired knowledge to novel and previously unencountered data.

Table I offers an insight into the model’s performance on previously unseen data, facilitating comparison with the performance of the ConvCfC model, as elaborated in Figure 4. These comparative findings underscore the models’ effective performance. Specifically, the sCCfC model achieves an average F1 score of 0.71 and an AUROC of 0.86, while the ConvCfC model attains an F1 score of 0.72 and an AUROC of 0.91 when assessed on the same dataset. These results emphasize the robust generalization capabilities of the sCCfC model, which closely aligns with the performance of the non-spiking ConvCfC model.

### E. Model Robustness

To thoroughly assess the model’s performance, we carried out supplementary evaluations utilizing the CPSC dataset. In these assessments, deliberate variations were introduced by selectively excluding specific channels from the 12-lead ECG data. This intentional manipulation of the input data evaluated the model’s robustness and ability to handle scenarios where input information is either incomplete or missing.

Figure 5 provides insights into the impact of omitted leads on the models’ F1 performance metric. As the number of removed leads increases, we observe a decrease in model performance. Notably, when comparing the performance of the sCCfC model to the ConvCfC model, it becomes apparent that while the initial performance of sCCfC might have been slightly lower, it exhibits greater resilience in the face of missing channels. As more channels are excluded from the data, the sCCfC model maintains its accuracy better than the ConvCfC model.

**FIG. 5.**
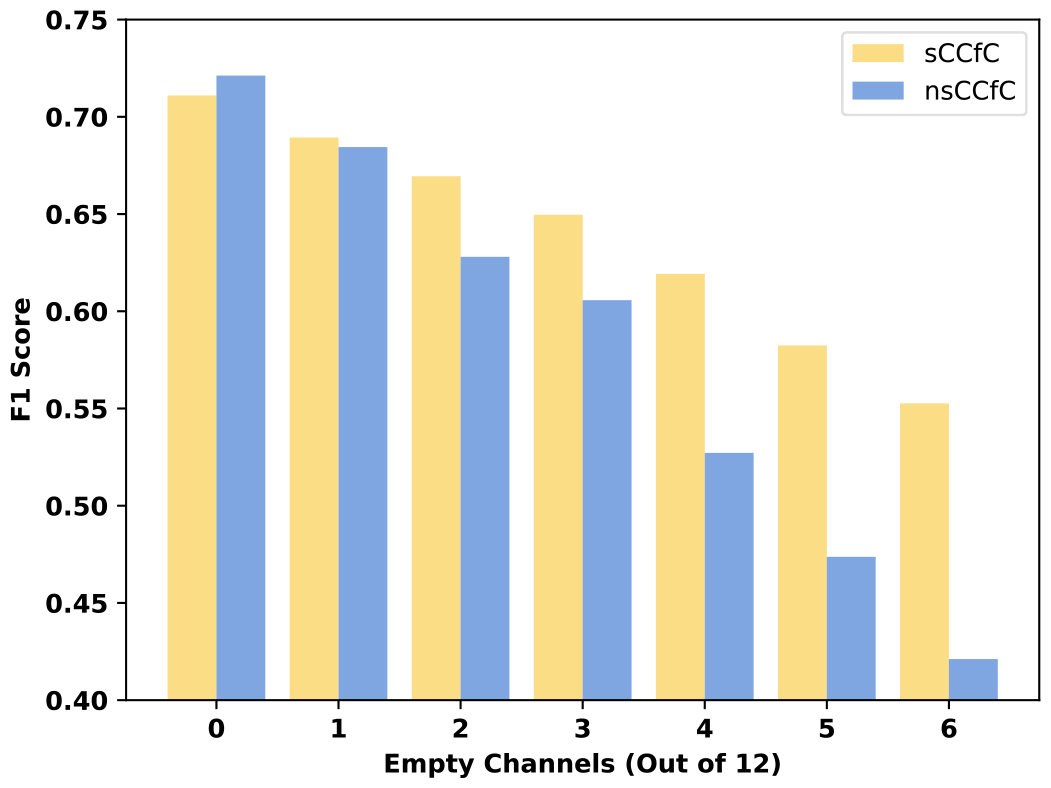
The F1 metrics of the models were examined about the presence of empty channels within the 12-lead CPSC ECG data.

### F. Model Performance with Single-Lead ECG

As a component of the study showcasing the model’s proficiency in handling single-lead ECG data, the model underwent training exclusively with Lead II ECG of the TNMG subset. The validation results of the model have been succinctly summarized in Table III.

**TABLE III.**
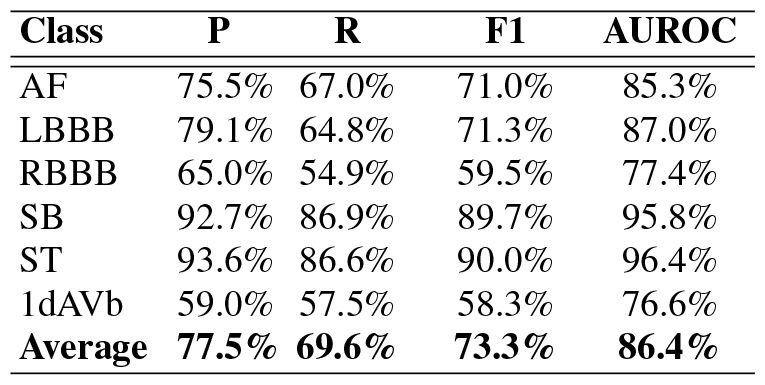
sCCfC Model Performance on Lead II ECG Data (P: Precision, R: Recall)

The model exhibits satisfactory performance, underscoring its capability to handle reduced-lead ECG data and signaling promising prospects for future applications.

## VII. DISCUSSION

The performance of the proposed sCCfC model on the TNMG subset dataset showcases its competence in classifying various cardiac abnormalities. The model’s precision, recall, F1-score, and AUROC values across different classes reflect its proficiency in distinguishing between normal and abnormal ECG patterns. Overall, sCCfC achieves an average F1-score of 0.81 and an average AUROC of 0.91. These metrics signify the model’s ability to effectively identify abnormal ECG patterns with a balanced trade-off between precision and recall.

In comparison, the ConvCfC model, which serves as a baseline, exhibits similar performance metrics. The average F1-score for ConvCfC is 0.83, and the average AUROC is 0.96. These results suggest that the sCCfC model, despite its spiking neural network architecture, maintains competitive performance with the non-spiking ConvCfC model. This finding highlights the potential of spiking neural networks in achieving efficient and effective ECG classification.

Table IV presents a comparative analysis of models transitioning to spiking neural networks, contrasting with similar works. It highlights that the proposed sCCfC has maintained its performance in conversion compared to other spiking neural networks.

**TABLE IV.**
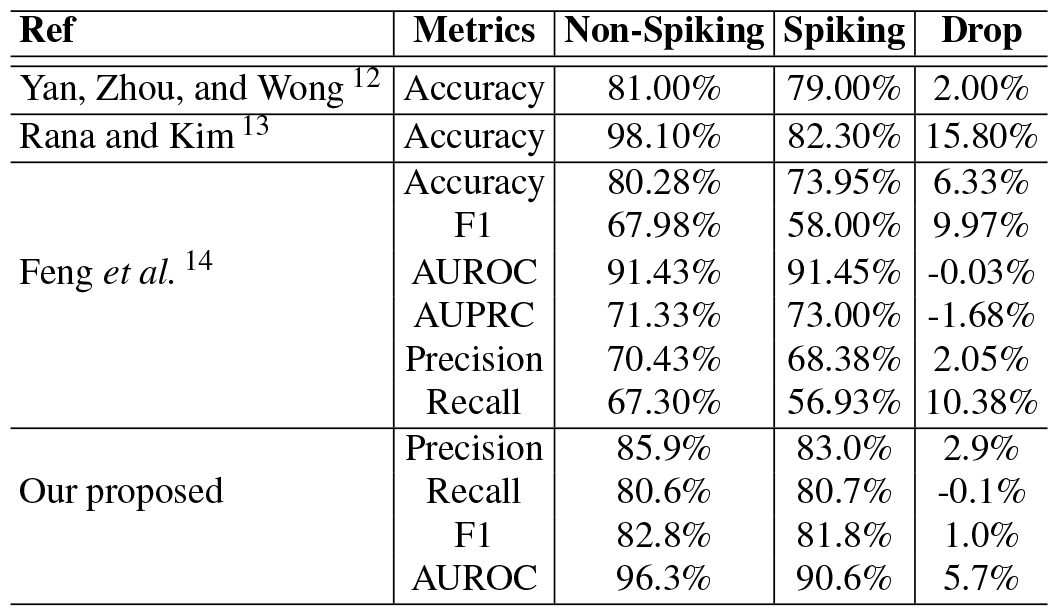
Previous Research on Normal Neural Networks Against Spiking Neural Networks.

One of the standout advantages of the sCCfC model is its significantly reduced estimated energy consumption compared to the ConvCfC model, owing to the inherent characteristics of Spiking Neural Networks (SNN). While the ConvCfC model consumes approximately 450 *µ*J/Inf when executed on a CPU, the sCCfC model demonstrates notably lower energy consumption, with only around 4.68 *µ*J/Inf when deployed on a neuromorphic chip like Loihi. This substantial decrease in energy consumption underscores the energy efficiency of SNNs, making them a suitable choice for deployment in battery-powered or energy-constrained devices. Moreover, the performance difference, as depicted in Figure 4, in comparison to other existing approaches shown in Table IV, exhibits a minimal drop in performance when transformed into SNNs.

The sCCfC model’s ability to undergo on-device finetuning on the Radxa Zero with performance improvements is noteworthy. This process demonstrates the model’s adaptability and resource efficiency, making it suitable for realworld applications requiring continuous learning and refinement. While this is a proof of concept demonstration conducted on a non-neuromorphic chip, it nonetheless showcases the model’s capabilities.

The evaluation of model generalization on the CPSC dataset, which contains ECG data with different abnormalities than the TNMG subset, provides valuable insights into the models’ adaptability to new and diverse data. The sCCfC model maintains competitive performance, with an average F1-score of 0.71 and an average AUROC of 0.86. While these metrics are slightly lower than those achieved on the TNMG subset, they indicate that the model can generalize effectively to new ECG abnormalities. When compared to the baseline model, the non-spiking ConvCfC model attained an average F1-score of 0.72 and an average AUROC of 0.89 on the CPSC dataset. These outcomes highlight that the sCCfC model continues to exhibit a robust capacity for transferring acquired knowledge to previously unencountered data, even when compared to its non-spiking counterpart.

The robustness of the sCCfC model is evident in its consistent performance across various levels of data incompleteness, where leads are progressively removed from the 12-lead ECG data. The model’s ability to maintain accuracy even when presented with partially available input information underscores its resilience and potential for handling noisy or incomplete data in clinical settings. In this domain, the sCCfC model has exhibited superior performance compared to the ConvCfC model, unveiling a captivating revelation.

## VIII. CONCLUSION

In conclusion, this research comprehensively analyses the sCCfC model for cardiac abnormality detection and diagnosis.

The sCCfC model demonstrated competitive performance metrics, closely matching the non-spiking ConvCfC model regarding average F1 score and AUROC. This suggests that spiking neural networks can achieve similar diagnostic accuracy while offering unique advantages.

The sCCfC model exhibited significantly lower energy consumption per inference than ConvCfC, making it a promising option for energy-efficient deployments, particularly when utilizing neuromorphic hardware like the Loihi chip.

The sCCfC model’s ability to undergo on-device finetuning on a single-board computer showcases its adaptability and resource efficiency, making it suitable for applications that require continuous learning and refinement.

The sCCfC model demonstrated strong generalization capabilities when applied to a novel dataset (CPSC) with different types of abnormalities. It maintained competitive diagnostic performance, emphasizing its potential for real-world applications.

The sCCfC model displayed resilience in handling missing channels in ECG data, outperforming the ConvCfC model as channels were omitted. This highlights the robustness of spiking neural networks to input variability.

The sCCfC model is promising for cardiac abnormality detection and diagnosis. Its combination of competitive performance, energy efficiency, adaptability, and robustness positions it as a valuable tool for healthcare professionals and researchers in cardiology.

Subsequent research and practical applications have the potential to leverage the advantages of spiking neural networks to propel the accuracy and efficiency of anomaly detection systems forward. In forthcoming endeavors, our focus will revolve around advancing on-chip learning, encompassing finetuning for personalization, augmenting memory optimization, and establishing corresponding testing protocols. Furthermore, we intend to broaden the capabilities of our potential portable device by integrating additional sensors. This approach will bring us closer to achieving real-time data training, thereby further enhancing the model’s adaptability and versatility.

## Data Availability

This research paper heavily depends on the publicly accessible CPSC dataset (http://2018.icbeb.org/Challenge.html) for its analysis and experimentation. Nevertheless, it is imperative to acknowledge that access to the TNMG dataset is inaccessible to the general public. To gain permission to access the TNMG dataset, one must formally request authorization from the data owner, and access can only be granted following approval. It is essential to emphasize that the TNMG dataset is not readily accessible to the wider public.

## IX. ACKNOWLEDGEMENT

Zhaojing Huang expresses gratitude for the support from the Australian Government’s Research Training Program (RTP).

## X. CODE AVAILABILITY

The code is available through the link https://github.com/NeuroSyd/Spiking-CfC. Please note that there may be specific terms, conditions, or usage restrictions associated with the code.

## SUPPLEMENTARY INFORMATION

### Neural circuit policies (NCP)

The synaptic connections among neurons in the NCP network constitute the foundational structure of the network. These synaptic links are established through various mechanisms:

1. Information transmission between consecutive layers involves source neurons transmitting data to target neurons via a specific number of synapses denoted as *n*_*so*_*−* _*t*_. These synapses’ distribution follows a Bernoulli distribution characterized by the probability *p*_2_^3^.
2. In cases where target neurons initially lack synapses, additional synapses (*m*_*so*_ *−* _*t*_) are introduced from source neurons. The quantity of these synapses is determined by a Binomial distribution with a probability denoted as *p*_3_. These synapses are randomly selected from an available pool of source neurons^3^.
3. Command neurons exhibit recurrent connections, forming synapses (*l*_*so*_*−* _*t*_) targeting command neurons. The establishment of these connections is governed by a Binomial distribution with a probability identified as *p*_4_^3^.

Together, these mechanisms collaboratively influence the flow of information and computational behaviors within the NCP model^3^.

Equation 2 illustrates the utilization of the semi-implicit Euler technique in the NCP model, where *I*_*in*_ denotes the ensemble of neurons acting as inputs to neuron *i*:

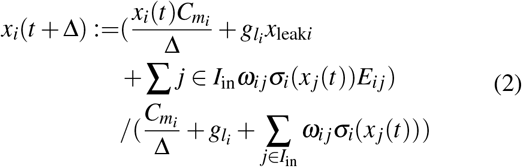

### Datasets

Our research involved a comprehensive evaluation of our proposed models using two distinct datasets. The first dataset, known as the CPSC dataset or the 12-lead ECG dataset, was originally created for The China Physiological Signal Challenge in 2018^20^. Its primary objective was to enable the automated detection of irregularities in both the rhythm and morphology of 12-lead ECGs. The second dataset utilized in our study is the Telehealth Network of Minas Gerais (TNMG) dataset^21^, which served as our primary training dataset for building and fine-tuning our models.

To thoroughly assess the effectiveness of our models, we utilized the CPSC dataset as a separate testing dataset. This approach enabled us to evaluate how well the models could generalize to new and unfamiliar data, which differed from the TNMG dataset.

Our study was specifically designed to gauge the models’ ability to generalize and perform effectively on unfamiliar data, following their training on the TNMG dataset. This evaluation holds paramount importance in establishing the reliability and practical utility of the models in real-world scenarios.

#### Telehealth Network of Minas Gerais (TNMG) Dataset

Our study utilized the TNMG dataset, comprising an extensive repository of 2,322,513 labeled instances of 12-lead electrocardiogram (ECG) data. Within this dataset, we encountered six distinct categories of abnormalities, namely Atrial Fibrillation (AF), Left Bundle Branch Block (LBBB), First Degree Atrioventricular Block (1dAVb), Right Bundle Branch Block (RBBB), Sinus Tachycardia (ST) and Sinus Bradycardia (SB)^21^. These original ECG recordings were sampled at a frequency of 400 Hz.

To construct a well-structured and balanced dataset for our model training, we adopted a systematic approach. We initiated this process by randomly selecting 3,000 data instances for each of the six distinct abnormalities. Additionally, we included 3,000 instances with no indications of abnormalities. Through this meticulous data selection process, we curated a comprehensive dataset, totaling 21,000 instances. In instances where patients exhibited multiple abnormalities, we randomly selected the remaining instances from the TNMG dataset to reach our specified subset size of 21,000.

The dataset underwent a meticulous normalization process, carefully adjusted to a uniform length of 4,096 readings. This thorough standardization not only guaranteed complete consistency but also significantly streamlined the data analysis and modeling procedures. Any readings surpassing this predetermined length were methodically removed, resulting in a more straightforward data processing pipeline and an improved ability to conduct meaningful comparisons. As illustrated in Table V, the resampled dataset exhibited a balanced gender distribution, emphasizing the significance of inclusivity and reinforcing the reliability of subsequent analyses. Additionally, the dataset closely resembled the age distribution observed in the broader population, further bolstering its appropriateness for research inquiries related to age.

**TABLE V.** The subset of TNMG data used in our study constitutes a carefully balanced dataset that includes six distinct abnormality categories. Furthermore, it demonstrates a greater representation of elderly patients, in line with the broader population demographics. The total incidences may exceed the total patients due to some patients exhibiting multiple abnormalities.

| **Breakdown** | **1dAVb** | **RBBB** | **LBBB** | **SB** | **AF** | **ST** | **Normal** |
| --- | --- | --- | --- | --- | --- | --- | --- |
| *Gender* |  |  |  |  |  |  |  |
| Female | 1477 | 2048 | 2109 | 1212 | 1639 | 2188 | 1862 |
| Male | 1757 | 2272 | 1334 | 2158 | 1922 | 1111 | 1138 |
| *Age* |  |  |  |  |  |  |  |
| 0-29 | 95 | 50 | 13 | 306 | 24 | 350 | 372 |
| 30-44 | 253 | 287 | 107 | 503 | 122 | 697 | 707 |
| 45-59 | 593 | 952 | 615 | 1071 | 418 | 920 | 963 |
| 60-74 | 1173 | 1740 | 1427 | 1072 | 1348 | 837 | 671 |
| 74-89 | 1007 | 1175 | 1144 | 393 | 1476 | 446 | 272 |
| 90+ | 113 | 116 | 137 | 25 | 173 | 49 | 15 |
| Total | 3234 | 4320 | 3443 | 3370 | 3561 | 3299 | 3000 |

Moreover, the adopted sampling strategy yielded a well-balanced distribution of diverse abnormalities, enabling a thorough examination of their attributes and consequences. This equilibrium in the dataset substantially enhanced the model’s learning process and overall performance, as detailed in the research conducted by^22^.

#### The dataset for the China Physiological Signal Challenge 2018 (CPSC)

The CPSC dataset comprises 12-lead electrocardiograms (ECGs) sampled at a frequency of 500 Hz. To ensure seamless compatibility with the CPSC dataset for training purposes, we meticulously adjusted the sampling rate of the TNMG data from its original 400 Hz to match the 500 Hz rate. This dataset is notably exceptional due to its inclusion of ECGs collected from patients diagnosed with a wide range of cardiovascular conditions and common rhythms. A team of expert annotators painstakingly labeled these ECGs, resulting in precise annotations for the identified abnormalities. In total, the dataset encompasses a comprehensive set of eight distinct types of abnormalities.

In a comprehensive evaluation of the model’s generalization abilities, we rigorously conducted tests employing four specific overlapping abnormalities meticulously chosen from the dataset: AF, RBBB, 1dAVb, and LBBB. It’s important to underscore that our study intentionally omits four other types of abnormalities: ST-segment Depression (STD), Premature Ventricular Contraction (PVC), Premature Atrial Contraction (PAC), and ST-segment Elevated (STE). The dataset is characterized and outlined in Table VI.

**TABLE VI.** The table offers an overview of the CPSC dataset, delineating the gender and age distribution for each of the abnormalities examined in this study. The total incidences may exceed the total patients due to some patients exhibiting multiple abnormalities.

| **Breakdown** | **1dAVb** | **RBBB** | **LBBB** | **AF** | **Normal** |
| --- | --- | --- | --- | --- | --- |
| *Gender* |  |  |  |  |  |
| Female | 232 | 653 | 119 | 530 | 1734 |
| Male | 490 | 1203 | 116 | 687 | 1321 |
| *Age* |  |  |  |  |  |
| 0-29 | 24 | 113 | 1 | 10 | 505 |
| 30-44 | 37 | 151 | 8 | 24 | 541 |
| 45-59 | 130 | 380 | 29 | 146 | 721 |
| 60-74 | 269 | 700 | 100 | 456 | 809 |
| 74-89 | 240 | 467 | 85 | 537 | 460 |
| 90+ | 22 | 45 | 12 | 44 | 19 |
| Total | 722 | 1856 | 235 | 1217 | 3055 |

In our data selection process, we took great care to exclude any entries in the dataset that contained missing readings. This meticulous approach resulted in a final dataset comprising 6,877 distinct ECG tracings. Following this, we standardized the data to ensure consistency and uniformity.

After conducting an extensive dataset analysis, we observed a gender disparity, wherein male patients were more prominently represented than their female counterparts. However, it’s important to highlight that the age distribution of the patients closely resembles that of the broader population, with a substantial proportion falling into older age groups. Nevertheless, a more detailed examination of abnormality distribution indicates a slight imbalance. Specifically, the occurrence of Left Bundle Branch Block (LBBB) is comparatively less frequent when contrasted with the prevalence of other abnormalities present in the dataset.

